# QBSafe: a Randomized Trial of a Novel Intervention to Improve Care for People Living With Type 2 Diabetes

**DOI:** 10.64898/2026.02.06.26345768

**Authors:** Kasia J Lipska, Megan E Branda, Anne W Camp, Mel Montosa, Rozalina G. McCoy, Victor Montori, Felipe Larios, Victor M. Montori

**Author notes:** Conferences: This work was presented at Academy Health, June 7-10, 2025, Minneapolis Convention Center, Minneapolis, MN and it will be presented at the Care That Fits 2025 conference, October 12, 2025, University of Paris Cite, Paris, France.

## Abstract

**Background:** Effective interventions are needed to support co-creation of diabetes care plans that fit patients’ lives. We evaluated the QBSafe agenda-setting kit (14 conversation cards) for its impact on care fit and glycemic control when added to usual primary care.

**Methods:** This single-center, clinician-level cluster-randomized, open-label trial was conducted at a federally qualified health center in New Haven, Connecticut (ClinicalTrials.gov NCT05553912). Clinicians and their patients with type 2 diabetes and HbA1c >8% were randomized 1:1 to usual care with or without QBSafe cards. In the intervention arm, patients selected up to 3 cards highlighting concerns about life with diabetes prior to their visit. Primary outcomes were change at 6 months in care fit (Illness Intrusiveness Ratings Scale, IIRS) and HbA1c, analyzed by intention to treat. Secondary outcomes were treatment burden (Treatment Burden Questionnaire, TBQ) and diabetes distress (Diabetes Distress Scale, DDS), and satisfaction with visits.

**Results:** Between February 2023 and July 2024, 143 participants (mean age 56 years; 61% female; 73% Hispanic; mean HbA1c 10%) were enrolled: 74 received usual care with QBSafe, 69 usual care alone. At 6 months, there were no significant between-arm differences in changes in IIRS (−3.9 [95% CI -10.4, 2.6]), HbA1c (−0.2% [95% CI -0.9, 0.5]), TBQ (1.0 [95% CI -16.6, 18.6]), or DDS (−0.1 [95% CI -0.4, 0.2]). Clinicians reported greater satisfaction when using QBSafe. Patient satisfaction was high and did not differ across arms.

**Conclusions:** QBSafe cards improved clinician satisfaction but did not improve care fit or glycemic control. Future tools should focus on helping clinicians respond effectively to patient-identified challenges.

## BACKGROUND

People living with type 2 diabetes are expected to manage complex daily routines, including dietary changes, physical activity, and medication regimens; self-monitor and respond to metabolic parameters; and coordinate access to ongoing clinical and laboratory assessments.^1^ These demands place a significant burden on patients and caregivers, who must dedicate substantial attention, time, and effort to their care.^2^ The daily lives of these individuals can thus be disrupted and constrained by these demands and by diabetes-related symptoms in those with hyperglycemia or hypoglycemia. These dual burdens – of illness and treatment – can reduce people’s quality of life and hinder their ability to attain their diabetes treatment goals and optimal health.^3^ In busy or under-resourced clinical settings, clinicians may overlook or deprioritize these challenges of living with diabetes, despite their importance to patients’ health and well-being.^4,5^ Ideally, patients and clinicians would engage in collaborative conversations in which these problematic situations are identified and addressed by co-creating plans of care that are maximally responsive to patient needs and priorities while being minimally disruptive of the lives of patients and caregivers.^6^

The best evidence to date demonstrates that clinicians and patients seldom discuss treatment burden and other patient-important issues that affect diabetes care.^7^ Many such issues remain unaddressed, as clinicians may prioritize intensifying therapies to improve measured outcomes, such as HbA1c, rather than working on making care fit. Effective interventions are needed to enable and support patients to voice both problems and successes in their ongoing care, thereby giving patients and clinicians an opportunity to collaborate on making care fit in the context of their lives.

The QBSafe agenda setting kit is a novel set of cards designed to help patients identify concerns, barriers, and successes (or points of pride) they would like to raise during their encounter with a clinician. A single-arm feasibility study evaluating the QBSafe prototype kit enrolling 7 clinicians and 84 patients found that 64% of patients found the cards helpful and 78% would recommend them to other patients.^8^ The cards also triggered collaborative conversations and, in a third of encounters, led to changes in the treatment program. However, whether this approach can meaningfully improve care and health outcomes among people with diabetes remained unexplored.

Here, we report the results of a larger, cluster-randomized, clinical trial examining the effects of adding a set of cards, the QBSafe agenda setting kit, to usual primary care on the outcomes of the fit of care and glycemic control. This study is focused on the evaluation of the QBSafe cards in the care of people with diabetes facing substantial socio-economic disadvantage and receiving care in a federally qualified health center. By exploring the impact of this intervention on both clinical and patient-reported outcomes, this study aims to contribute to the development of scalable strategies for improving diabetes care and enhancing patient and clinician experiences.

## METHODS

### Study design

This clinician-level cluster randomized trial compared the effect of usual primary care with and without QBSafe agenda setting kit (ASK) cards on fit of care and glycemic control for adults with type 2 diabetes. The intervention was designed to support patients and their clinicians in identifying aspects of living with type 2 diabetes and its treatment in order to set the agenda for problem-solving conversations during the clinical encounter.^8,9^ The trial was registered at ClinicalTrials.gov (NCT05553912) and the protocol adhered to standard SPIRIT recommendations.^10,11^ Reporting adhered to the CONSORT guidelines for cluster randomized trials to allow for assessment of the trial soundness.^12,13^ All study procedures were reviewed and approved by the Institutional Review Board at Yale University.

### Setting

The trial took place at the Fair Haven Community Health Care (FHCHC), a federally qualified health center (FQHC) that cares for a diverse, bilingual (Spanish and English) population comprised of over 21,000 patients, of which more than 1,500 patients live with diabetes. We chose to test QBSafe at FHCHC because the center cares for a high-risk patient population with multiple medical and socioeconomic barriers to health, and thus is likely to benefit from holistic interventions to improve diabetes management and to align care with complex life realities. FHCHC serves a predominantly racial and ethnic minority population (with a high burden of diabetes), younger in age than the general diabetes population (with a high burden of complications^14^), and with a high prevalence of actionable social risk factors, such as food insecurity, housing instability, difficulties with transportation or paying bills,^15^ that may reduce their capacity to manage diabetes. Each of these factors make patients in this care setting likely to need and potentially benefit from interventions to achieve care that fits within their contexts.^16^

### Participants

Eligible clinicians were any primary care physicians, nurse practitioners, and physician assistants who participate in the care of patients with type 2 diabetes and prescribe medications for them. Eligible clinicians were recruited after a presentation of the trial at their regularly scheduled staff meeting. All clinicians provided written informed consent to participate in the trial prior to the enrollment of their first patient.

Eligible patients were adults (age 18 years or older), diagnosed with type 2 diabetes, able to sign informed consent, fluent in either English or Spanish, and with an HbA1c >8% for whom lower glycemic control levels were deemed desirable by their clinician. Clinicians reviewed the list of potentially eligible participants and identified patients for whom a HbA1c >8% was a sensible target (e.g., because of limited life expectancy) and these patients were not eligible to participate in the trial.

A trained study coordinator identified eligible patients through appointment lists and recruited them via phone (asking patients to arrive early to their scheduled appointment to complete the consent process), or in person at the time of their scheduled visit. All patients included in the trial provided written informed consent. In the spirit of minimally disruptive research, all study activities occurred within scheduled appointments, avoiding the need for additional research-only visits.

Data from the medical record were abstracted for all enrolled patients to capture demographic characteristics (including sex, race, ethnicity, federal poverty level category), clinical comorbidities (using ICD-10 codes from the problem list and outpatient encounters), and medication prescription and fill (dispensing) data. The time frame for collection was 2 years prior to enrollment to 6 months post enrollment, allowing for a window of 4 to 8 months post enrollment for outcome collection.

Clinician characteristics were collected via a one-time survey at time of consent (age, sex [female, male], race, ethnicity, type of clinician [physician on staff, nurse practitioner, physician assistant], fluency in Spanish vs use of interpreter services).

### Randomization and Masking

Clinicians were randomized in a 1:1 ratio by the study statistician using the Pocock-Simon method to balance clinician characteristics between the QBSafe arm and the usual care arm. Minimization was based on clinician site and Spanish language proficiency (fluent or requiring an interpreter). The study coordinator assessed patient eligibility and enrolled patient participants using the REmote Data Capture (REDCap) system, with assignment based on their treating clinician. Except for patients and clinicians, who were informed that the trial tested different ways of conducting diabetes care encounters and were blind to the specific objectives and hypotheses of the trial, all study personnel were able to discern participant allocation.

### Procedures

In the intervention group, the study coordinator handed patients the set of 14 cards (QBSafe Agenda Setting Kit, https://patientrevolution.org/qbsafe, *Supplement*) as they waited for their clinician either in the waiting room or in the exam room. They were asked to select 0 to 3 cards that best fit their situation and to bring them up at the beginning of the consultation. Clinicians randomized to the QBSafe intervention were asked to conduct the encounter as usual but to review and respond to the cards first. Prior to their first visit with a study patient, they received a brief training on the use of QBSafe cards (in small groups or 1:1). During training, clinicians had the opportunity to try the QBSafe cards and to access materials with suggested responses (*Supplement*). All resources for responses were also available online, via brief video response examples.

In the control group, clinicians were asked to conduct their encounter as usual. To limit contamination, we did not give access to the QBSafe cards to clinicians allocated to the control arm, a feature, along with clinician training, that justified our choice of using a clustered randomized design.

### Primary patient-reported outcome

Patient-reported outcomes were captured using validated versions of the instruments in English and Spanish, collected at baseline and 6 months later, allowing for a window of 4-8 months. The primary patient-reported outcome (care that fits) was the change in illness and treatment intrusiveness measured by the validated Illness Intrusiveness Ratings Scale (IIRS).^3^ IIRS comprises of 13 items that ask patients to rate the degree to which their “illness and/or its treatment” interfere with life domains central to quality of life (health, diet, work, active and passive recreation, financial situation, relationships, sex life, family, and others). Thus, IIRS provides a direct measure of the extent to which disease and/or treatment interfere with valued lifestyles, activities, and interests. Existing evidence demonstrates IIRS’s reliability, validity, and sensitivity to change.^3^ This measure was selected as the primary outcome (instead of diabetes distress or treatment burden) because it better aligns with the conceptual model (fit of care) and captures the sum of illness- and treatment- burden.

### Primary clinical outcome

The primary clinical outcome, a measure of glycemic control, was the change in HbA1c from baseline (obtained prior to the index encounter) to 6 months (obtained as close as possible to this time point) after the index encounter.

### Secondary outcomes

Since IIRS is not a diabetes-specific measure, we also evaluated several diabetes-specific secondary outcomes.

Diabetes distress was measured by the validated Diabetes Distress Scale (DDS-17).^17^ Diabetes distress refers to “the worries, concerns, fears and threats that are associated with struggling with a demanding chronic disease like diabetes over time, including its management, threats of complications, potential loss of functioning and concerns about access to care.”^18^ Total and subscale scores are calculated using mean item scores, which are then categorized as little or no distress (< 2.0), moderate distress (≥ 2.0 and ≤ 2.9) and high distress (≥3.0). Moderate and high distress are considered ‘clinically significant.’^19^

Since IIRS combines the burdens of illness and treatment, we sought to isolate the potential impact of the intervention on the burden of treatment alone via the Treatment Burden Questionnaire (TBQ).^20^ The TBQ is composed of 15 items rated on a Likert-type scale ranging from 0 (no impact), to 10 (considerable impact). It assesses the burden associated with taking medicine, self-monitoring, laboratory tests, doctor visits, need for organization, administrative tasks, following advice on diet and physical activity, costs of treatment, and social impact of the treatment. Item scores can be summed into a global score, ranging from 0 to 150.^21^ It is both valid and reliable.^20^

### Other outcome measures

Health literacy was assessed with a single screening question about confidence in filling out medical forms.^22,23^ In clinical settings, the instrument allows clinicians to appropriately adapt their communication practices to the patient’s health literacy level.

Self-management was measured using the Summary of Diabetes Self-Care Activities (SDSCA).^24^ SDSCA is a brief self-reported questionnaire assessing the following aspects of the diabetes regimen: general diet, specific diet, exercise, blood-glucose testing, foot care, and smoking. The measure has been demonstrated to be both valid and reliable.

To assess global quality of life, we used the 0 (worst possible)-to-10 (best possible) single-item visual analogue scale, which is a valid, reliable, and responsive measure recommended for use in clinical trials.^25^

Patient satisfaction with the intervention was assessed with a single-item question on a 5-point Likert scale that addresses whether they would recommend the approach used during their encounter (which could have been either usual care alone or with the QBSafe ASK) to other patients. This is a responsive measure used during validation of the QBSafe ASK and used in prior trials of shared decision-making interventions.^26^ The quality of communication was assessed with a modified version of 3 questions from the CAHPS Clinician and Group survey.^27^ These questions indicate the extent to which communication is patient-centered, covering technical (explain things in a way you could understand) and effective (show respect for what you have to say) communication.

Clinicians rated their satisfaction with the approach used during the encounter (which could have been usual care alone or usual care with the QBSafe ASK) on a 5-point Likert-type scale. They also rated, on a 5-point Likert-type scale, how confident they felt in responding to patient concerns. Both are responsive measures we used during the validation of the QBSafe ASK and have used in prior trials of shared decision-making interventions.^28^

Finally, medical and pharmacy records were abstracted to assess the doses and types of diabetes medications prior to encounter, immediately post encounter, and 6 months post encounter. We calculated adherence to diabetes medications based on the proportion of days covered (PDC) 6 months prior to and 6 months post encounter (available in Epic based on pharmacy data), defining high adherence as PDC ≥80%, intermediate adherence as PDC of 40%-79%, and low adherence as PDC <40%.^29,30^ For those on multiple medications, we used the average PDC across their treatment. Patients who started a new medication post baseline do not have a PDC reported and not all patients had a complete 6-month follow-up in the EHR for PDC. We also assessed healthcare utilization: any hospitalizations or emergency department (ED) visits within 6 months post encounter.

### Statistical Analysis

With a target sample size of 10 clinicians and their 130 patients (65 per arm), we estimated being able to detect a mean between-arm difference of 7.5 points on IIRS (primary outcome) and 77% and 99% power to detect a between-arm difference in HbA1c of 0.3 and 0.6%, respectively. To account for potential attrition, the sample size was increased by 20% to 155 patients.

Baseline clinicians and patient characteristics were summarized with continuous values being reported as means and standard deviations and categorical values reported as counts and frequencies. Baseline characteristics were assessed for clinical and statistical differences, which were accounted for in regression models that adjusted for intervention arm as well. As the clinician is a clustering effect, a hierarchical generalized linear model (HGLM) was conducted to assess each outcome of interest. For analysis of the outcomes collected prior to visit and at 6 months post enrollment (IIRS, DDS-17, TBQ, quality of life, HbA1c, SDSCA, medication adherence, and healthcare utilization), the 6-month response/value was modeled adjusting for the baseline value, arm of interest and random effect of clinician. If the baseline value was missing, a dummy variable was included in the model to indicate missing or not so that the patient’s response could still be assessed. Mean difference between arms with a 95% confidence interval was reported.

Next, we explored potential interactions between the intervention and pre-specified patient factors (age, sex, health literacy, and language) within outcomes of IIRS and A1c. While we were not powered to test for these interactions given the size of the trial, we explored any trends that should be accounted for in further work with a sufficient sample size. To conduct this analysis, the models were extended using interaction terms to examine several types of heterogeneity of treatment effect due to possible changes or improvements in effectiveness of the intervention (interaction of intervention with patient characteristic).

## RESULTS

### Baseline Characteristics

Between February 16, 2023, and July 26, 2024, we assessed 888 patients for inclusion. **Figure 1** depicts the flow of clinicians and patients as trial participants. Among 12 eligible clinicians who were approached to participate in the study, 11 agreed to participate, 6 were randomly allocated to QBSafe and 5 to usual care arm. For the QBSafe arm, 471 potentially eligible patients of the 6 clinicians were screened, and we enrolled 83 patients. After enrollment, the most recent HbA1c excluded 9 patients who became ineligible as their latest HbA1c was ≤8%, thus leaving 74 patients in the QBSafe arm. For the usual care arm, 417 potentially eligible patients of the 5 clinicians were screened, and we enrolled 72. After enrollment, the most recent HbA1c excluded 3, leaving 69 patients in the usual care arm.

**Figure 1:**
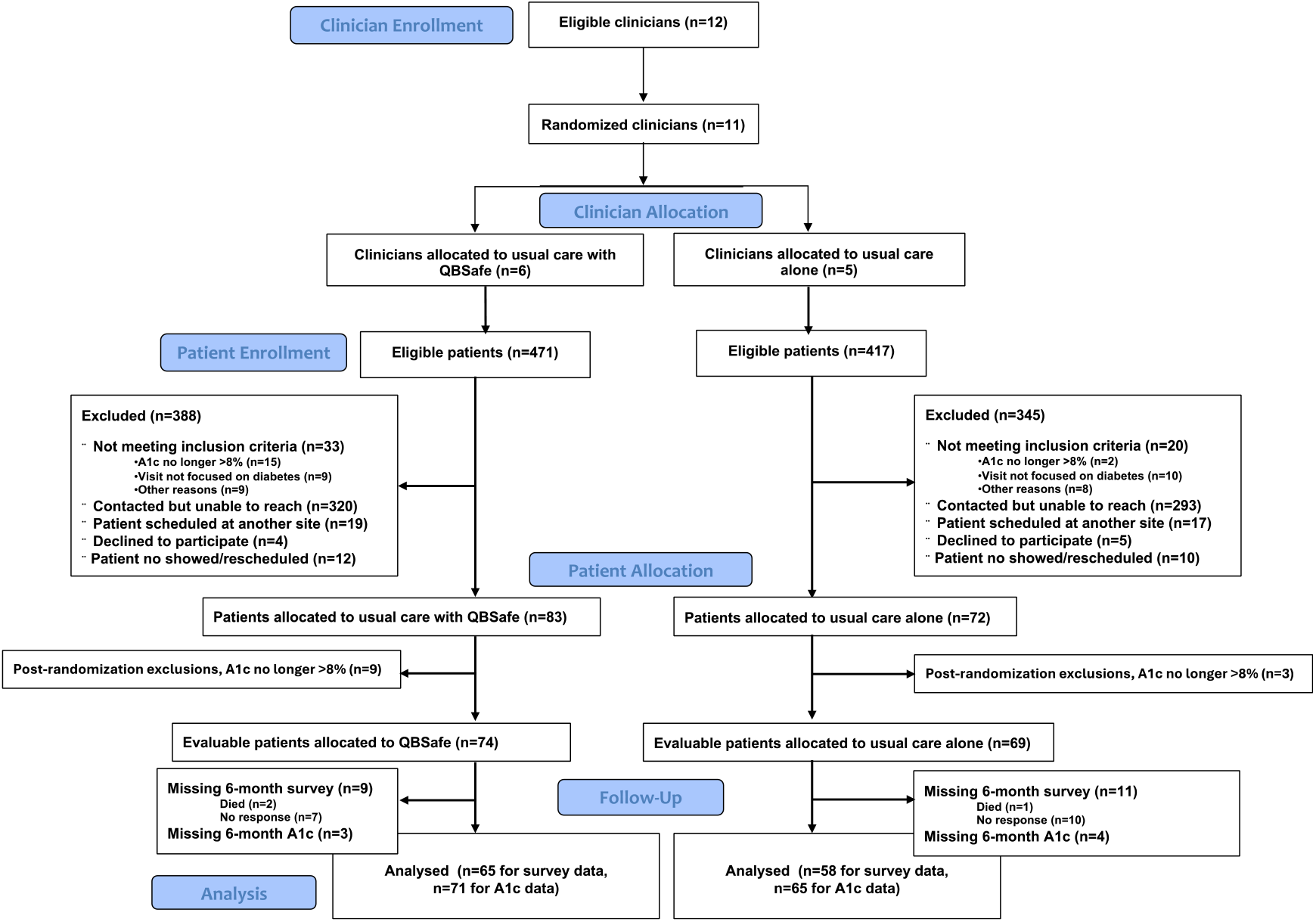
CONSORT diagram

In the QBSafe arm there were 3 male and 3 female clinicians including 4 physicians and 2 nurse practitioners. Three were fluent in Spanish. In the usual care arm, there were 2 male and 3 female clinicians, with 4 physicians and 1 nurse practitioner. Two were fluent in Spanish.

**Table 1** describes the baseline characteristics of the 143 participating patients: mean age was 56 (SD 12) years, 61% were female, 73% Hispanic, and 62% reported Spanish as their primary language. Among these patients, 91% reported family income below 150% of the federal poverty level, 37% were covered by Medicaid and 25% were uninsured (self-pay). A quarter of patients had established retinopathy and 31% had neuropathy. Mean baseline HbA1c level was 10% (SD 1.7), with 43% of participants starting the trial with a HbA1c ≥ 10%.

**Table 1:** Characteristics of patient participants in the QBSafe trial by study arm.

|  | Study Arm<br>Usual Care<br>(N=69) | QBSafe<br>(N=74) | Total<br>(N=143) | ICC |
| --- | --- | --- | --- | --- |
| **Age, Mean (SD)** | 53.8 (10.9) | 58.4 (12.3) | 56.2 (11.8) | 0.20 |
| **Female Sex, n (%)** | 44 (63.8%) | 43 (58.1%) | 87 (60.8%) | 0.07 |
| **Race, n (%)** |  |  |  |  |
| White | 19 (27.5%) | 32 (43.2%) | 51 (35.7%) | 0.005 |
| Black | 15 (21.7%) | 13 (17.6%) | 28 (19.6%) | 0.005 |
| More than one race | 0 (0.0%) | 1 (1.4%) | 1 (0.7%) | 0 |
| Other | 29 (42.0%) | 24 (32.4%) | 53 (37.1%) | 0.02 |
| Unknown | 6 (8.7%) | 4 (5.4%) | 10 (7.0%) | 0 |
| **Hispanic or Latino Ethnicity, n (%)** | 53 (76.8%) | 51 (68.9%) | 104 (72.7%) | 0.02 |
| **Primary Language, n (%)** |  |  |  | 0.01 |
| English | 21 (30.4%) | 34 (45.9%) | 55 (38.5%) |  |
| Spanish | 48 (69.6%) | 40 (54.1%) | 88 (61.5%) |  |
| **Was an interpreter present?, n (%)** |  |  |  | 0.62 |
| Yes, on the phone | 14 (20.3%) | 8 (10.8%) | 22 (15.4%) |  |
| Yes, in person | 2 (2.9%) | 5 (6.8%) | 7 (4.9%) |  |
| No | 53 (76.8%) | 61 (82.4%) | 114 (79.7%) |  |
| **Medical Insurance, n (%)** |  |  |  |  |
| Commercial | 9 (13.0%) | 11 (14.9%) | 20 (14.0%) | 0 |
| Medicaid | 27 (39.1%) | 26 (35.1%) | 53 (37.1%) | 0.02 |
| Medicare | 13 (18.8%) | 22 (29.7%) | 35 (24.5%) | 0.23 |
| Self-Pay/Uninsured | 20 (29.0%) | 15 (20.3%) | 35 (24.5%) | 0.12 |
| **Federal Poverty Level, n (%)** |  |  |  | 0 |
| 0-150% | 55 (90.2%) | 61 (92.4%) | 116 (91.3%) |  |
| >150% | 6 (9.8%) | 5 (7.6%) | 11 (8.7%) |  |
| Missing | 8 | 8 | 16 |  |
| **Confidence in Filling Out Medical Forms, n (%)** |  |  |  | 0 |
| Not at all/A little bit confident | 5 (7.4%) | 10 (13.5%) | 15 (10.5%) |  |
| Somewhat confident | 7 (10.3%) | 9 (12.2%) | 16 (11.3%) |  |
| Quite a bit/Extremely confident | 56 (82.4%) | 55 (74.3%) | 111 (77.6%) |  |
| **Comorbidities, n (%)** |  |  |  |  |
| Cardiovascular disease | 5 (7.2%) | 12 (16.2%) | 17 (11.9%) | 0 |
| Cerebrovascular disease | 3 (4.3%) | 5 (6.8%) | 8 (5.6%) | 0 |
| Hyperlipidemia | 52 (75.4%) | 55 (74.3%) | 107 (74.8%) | 0.24 |
| Hypothyroidism | 5 (7.2%) | 6 (8.1%) | 11 (7.7%) | 0 |
| **Diabetes Complications, n (%)** |  |  |  |  |
| Nephropathy | 11 (15.9%) | 13 (17.6%) | 24 (16.8%) | 0 |
| Neuropathy | 19 (27.5%) | 25 (33.8%) | 44 (30.8%) | 0.07 |
| Retinopathy | 17 (24.6%) | 18 (24.3%) | 35 (24.5%) | 0 |
| **Baseline Diabetes Medications** |  |  |  |  |
| Count of Diabetes Medications, n (SD) | 2.8 (1.0) | 2.8 (0.9) | 2.8 (1.0) | 0.08 |
| Metformin, n (%) | 55 (79.7%) | 59 (79.7%) | 114 (79.7%) | 0.21 |
| Sulfonylureas, n (%) | 16 (23.2%) | 19 (25.7%) | 35 (24.5%) | 0.05 |
| GLP-1, n (%) | 21 (30.4%) | 28 (37.8%) | 49 (34.3%) | 0.0 |
| SGLT-2 Inhibitors, n (%) | 36 (52.2%) | 35 (47.3%) | 70 (49.0%) | 0.05 |
| DPP-4 inhibitors, n (%) | 20 (29.0%) | 20 (27.0%) | 39 (27.3%) | 0.01 |
| TZDs, n (%) | 1 (1.4%) | 2 (2.7%) | 3 (2.1%) | 0.53 |
| Meglitinides, n (%) | 0 (0.0%) | 1 (1.4%) | 1 (0.7%) | 0.0 |
| Insulin, n (%) | 41 (59.4%) | 43 (58.1%) | 84 (58.7%) | 0.002 |
| Percent of Days Covered (Adherence), % (SD) | 67.2 (31.6) | 70.6 (28.3) | 69.0 (29.8) | 0.002 |
| **BMI, Mean (SD)** | **31.7 (5.12)** | **30.9 (5.63)** | **31.3 (5.39)** | **0.04** |
| **BMI categories, n (%)** |  |  |  |  |
| 18.5 to 24.9 | 6 (8.7%) | 8 (10.8%) | 14 (9.8%) |  |
| 25 to 29.9 | 21 (30.4%) | 24 (32.4%) | 45 (31.5%) |  |
| 30.0 to 39.9 | 36 (52.2%) | 38 (51.4%) | 74 (51.7%) |  |
| 40.0+ | 6 (8.7%) | 4 (5.4%) | 10 (7.0%) |  |
| **HbA1c - Baseline, Mean (SD)** | **10.2 (1.76)** | **9.9 (1.70)** | **10.0 (1.73)** | **0.001** |
| **HbA1c categories, n (%)** |  |  |  |  |
| 8.0 to <9.0 | 18 (26.1%) | 28 (37.8%) | 46 (32.2%) |  |
| 9.0 to < 10.0 | 22 (31.9%) | 16 (21.6%) | 38 (26.6%) |  |
| 10.0+ | 29 (42.0%) | 30 (40.5%) | 59 (41.3%) |  |
| **Baseline Patient Reported Outcome (PRO) Scales** |  |  |  |  |
| Illness Intrusiveness Score, Mean (SD) | 36.6 (17.52) | 34.7 (15.68) | 35.6 (16.56) | 0.02 |
| IIRS subscale: Relationship and personal, Mean (SD) | 2.2 (1.58) | 2.2 (1.40) | 2.2 (1.49) | 0.05 |
| IIRS subscale: Intimacy, Mean (SD) | 2.7 (2.15) | 2.2 (1.82) | 2.4 (1.99) | 0.002 |
| IIRS subscale: Instrumental, Mean (SD) | 3.3 (1.64) | 3.1 (1.56) | 3.2 (1.60) | 0.01 |
| Diabetes Distress Scale - Overall, Mean (SD) | 2.5 (1.07) | 2.2 (1.02) | 2.3 (1.05) | 0.03 |
| DDS: Physician-related Distress, Mean (SD) | 1.8 (0.92) | 1.6 (0.75) | 1.7 (0.84) | 0.04 |
| DDS: Emotional Burden, Mean (SD) | 3.1 (1.55) | 2.7 (1.50) | 2.9 (1.53) | 0.04 |
| DDS: Regimen-related Distress, Mean (SD) | 3.0 (1.54) | 2.6 (1.35) | 2.8 (1.45) | 0.04 |
| DDS: Interpersonal Distress, Mean (SD) | 2.0 (1.29) | 2.0 (1.44) | 2.0 (1.36) | 0.04 |
| Treatment Burden Questionnaire, Mean (SD) | 45.6 (38.26) | 37.8 (35.37) | 41.5 (36.87) | 0.08 |
| QOL, Mean (SD) | 6.2 (2.8) | 6.2 (2.9) | 6.2 (2.9) | 0.01 |

At baseline, mean IIRS score was 35.6 (SD 16). Although normative levels of IIRS have not been established, these levels of illness intrusiveness are comparable to those reported by patients living with cancer or after organ transplantation.^3^ Diabetes distress as measured by DDS demonstrated a mean score of 2.3, consistent with moderate distress,^17^ with highest levels of distress on the emotional and regimen-related subscales.

### Patient Reported Outcome Measures

At 6 months of follow-up, 65/74 (88%) of QBSafe arm and 58/69 (84%) usual care arm patients had available follow-up survey data. After adjusting for clustering and baseline values, there were no significant differences between arms in changes in IIRS (−3.9, 95% CI -10.4, 2.6), DDS (−0.09, 95% CI -0.4, 0.2), TBQ (1.01, 95% CI -16.6, 18.6), or quality of life (−0.53, 95% CI - 0.78,1.84) from baseline to 6 months (**Table 2**). There were also no significant differences between arms in changes in the Summary of Diabetes Self-Care Activities (*Supplement*).

**Table 2:** QBSafe trial patient reported and clinical outcomes at 6 months by study arm.

|  | Study Arm<sup>3</sup> |  | DID (95% CI) | P-value | ICC |
| --- | --- | --- | --- | --- | --- |
|  | Usual Care<br>(N=69) | QBSafe<br>(N=74) |  |  |  |
| Patient Reported Outcomes<sup>3</sup> |  |  |  |  |  |
| Illness Intrusiveness Score | 37.1 (18.7) | 32.1 (15.1) | -3.9 (-10.4, 2.6) | 0.24<sup>1</sup> | 0.05 |
| Illness Intrusiveness Subscales: |  |  |  |  |  |
| Relationship and personal | 2.4 (1.7) | 2.1 (1.4) | -0.3 (-1.1, 0.5) | 0.44<sup>1</sup> | 0.09 |
| Intimacy | 2.4 (1.8) | 2.0 (1.8) | -0.3 (-0.8, 0.3) | 0.39<sup>1</sup> | 0.02 |
| Instrumental | 3.4 (1.7) | 3.0 (1.5) | -0.3 (-0.8, 0.2) | 0.28<sup>1</sup> | 0.01 |
| Diabetes Distress Scale | 2.3 (1.0) | 2.0 (1.0) | -0.09 (-0.4, 0.2) | 0.49<sup>1</sup> | 0.03 |
| Diabetes Distress Subscales: |  |  |  |  |  |
| Emotional Burden | 2.9 (1.5) | 2.4 (1.4) | -0.2 (-0.5, 0.2) | 0.12<sup>1</sup> | 0.02 |
| Physician-related Distress | 1.6 (0.6) | 1.5 (0.8) | -0.1 (-0.3, 0.1) | 0.45<sup>1</sup> | 0.03 |
| Regimen-related Distress | 2.7 (1.4) | 2.3 (1.3) | -0.2 (-0.6, 0.2) | 0.32<sup>1</sup> | 0.04 |
| Interpersonal Distress | 1.9 (1.3) | 1.8 (1.3) | -0.04 (-0.4, 0.3) | 0.82<sup>1</sup> | 0.06 |
| Treatment Burden Questionnaire | 46.8 (44.4) | 41.2 (44.8) | 1.01 (-16.6, 18.6) | 0.91<sup>1</sup> | 0.11 |
| TBQ <=59 | 40 (70.2%) | 46 (69.7%) |  |  |  |
| > 59 | 17 (29.8%) | 20 (30.3%) |  |  |  |
| Quality of life | 6.4 (2.8) | 7.0 (2.6) | -0.53 (-0.78,1.84) | 0.42<sup>1</sup> | 0.10 |
| Clinical Outcomes |  |  |  |  |  |
| HbA1c<sup>4</sup> | 9.1 (1.8) | 8.8 (1.8) | -0.2 (-0.9, 0.5) | 0.59<sup>1</sup> | 0.04 |
| HbA1c categories, n (%) |  |  |  | 0.47<sup>2</sup> |  |
| <8.0 | 17 (26.2%) | 26 (36.6%) |  |  |  |
| 8.0 to <9.0 | 14 (21.5%) | 16 (22.5%) |  |  |  |
| 9.0 to < 10.0 | 14 (21.5%) | 10 (14.1%) |  |  |  |
| 10.0+ | 20 (30.8%) | 19 (26.8%) |  |  |  |
| Medications |  |  |  |  |  |
| Medication Count post encounter | 2.8 (1.1) | 2.9 (0.9) | 0 (-0.2, 0.2) | 0.99<sup>1</sup> | 0.06 |
| New Medications 1+ | 16 (23.2%) | 15 (20.3%) |  | 0.68<sup>2</sup> | 0.01 |
| Change in existing medication 1+ | 16 (23.2%) | 25 (33.8%) |  | 0.04<sup>2</sup> | 0.01 |
| Stopping a medication 1+ | 8 (11.6%) | 7 (9.5%) |  | 0.62<sup>2</sup> | 0.03 |
| Medication count at 6 months | 2.9 (1.1) | 3.0 (1.0) | 0.03 (-0.3, 0.3) | 0.84<sup>1</sup> | 0.006 |
| New medication 1+ | 7 (10.1%) | 10 (13.5%) |  | 0.14<sup>2</sup> | 0.05 |
| Change in existing medication 1+ | 3 (4.3%) | 7 (9.5%) |  | 0.07<sup>2</sup> | 0.005 |
| Stopping a medication 1+ | 7 (10.1%) | 8 (10.8%) |  | 0.87<sup>2</sup> | 0.02 |
| Percent of days covered<sup>5</sup> | 64.6 (29.9) | 77.2 (27.7) | 11.7 (1.3, 22.2) | 0.03<sup>1</sup> | 0.08 |

<sup>1</sup>Mixed generalized linear model adjusting for a fixed effect of arm and baseline score for the scale, and a random effect of clinician;
<sup>2</sup>Rao-Scott Chi-Square p-value – accounting for clustering of clinician, no baseline scale adjustment;
<sup>3</sup>Missing N=20 (Usual Care N=12, QBSafe N=8);
<sup>4</sup>Missing N=7 (Usual Care N=4, QBSafe N=3);
<sup>5</sup>Missing All PDC values Usual Care N=1;

### Clinical Outcomes

At 6 months of follow-up, 71/74 of QBSafe arm and 65/69 of usual care arm patients had available HbA1c data. After adjusting for clustering and baseline values, there were no significant differences between arms in changes in HbA1c (−0.2, 95% CI, -0.9, 0.5) from baseline to 6 months (**Table 2**). At 6 months, 27% of QBSafe and 31% of usual care participants had HbA1c ≥10%, while 37% of QBSafe and 26% of usual care participants had A1c levels ≤8%.

There were no differences between arms in the number of diabetes medications used, or in additions or discontinuations of diabetes medications (**Table 2**). However, post-encounter changes in medications (such as dose, frequency, or a change of medication within the same class) were higher in the QBSafe arm vs. usual care (34% vs. 23%, p=0.04) and adherence to diabetes medications as measured by the proportion of days covered was significantly higher in the QBSafe arm compared with usual care at 6 months (77.2% vs. 64.6%, mean difference 11.7% [95% CI 1.3, 22.2]) (**Table 2**).

At 6 months, 16 (24%) usual care patients and 13 (18%) QBSafe patients had an emergency department visit; 12 (18%) usual care patients and 10 (14%) patients in the QBSafe arm were hospitalized. There were no significant differences across study arms.

### Participant Satisfaction

Clinicians randomized to QBSafe arm were more likely to report feeling satisfied with the way the visit went and feeling confident in responding to patient concerns compared with clinicians randomized to usual care (**Table 3**). Patients’ levels of satisfaction with the approach used during the encounter and communication with their clinicians were overall very high and did not vary across arms (**Table 3**).

**Table 3:**
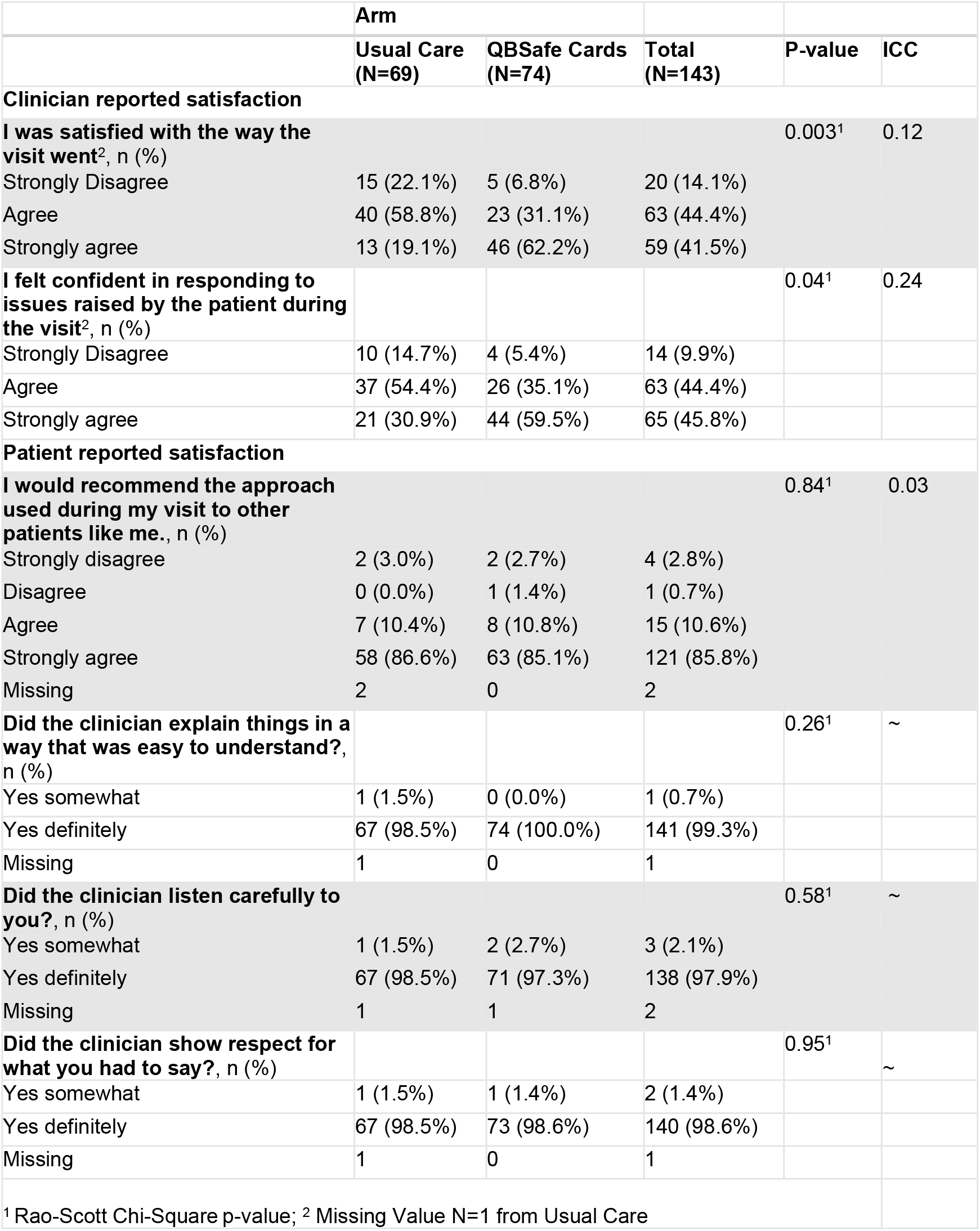
QBSafe trial participant satisfaction outcomes by trial arm.

|  | Arm |  |  |  |  |
| --- | --- | --- | --- | --- | --- |
|  | Usual Care<br>(N=69) | QBSafe Cards<br>(N=74) | Total<br>(N=143) | P-value | ICC |
| **Clinician reported satisfaction** |  |  |  |  |  |
| **I was satisfied with the way the visit went<sup>2</sup>, n (%)** |  |  |  | 0.003<sup>1</sup> | 0.12 |
| Strongly Disagree | 15 (22.1%) | 5 (6.8%) | 20 (14.1%) |  |  |
| Agree | 40 (58.8%) | 23 (31.1%) | 63 (44.4%) |  |  |
| Strongly agree | 13 (19.1%) | 46 (62.2%) | 59 (41.5%) |  |  |
| **I felt confident in responding to issues raised by the patient during the visit<sup>2</sup>, n (%)** |  |  |  | 0.04<sup>1</sup> | 0.24 |
| Strongly Disagree | 10 (14.7%) | 4 (5.4%) | 14 (9.9%) |  |  |
| Agree | 37 (54.4%) | 26 (35.1%) | 63 (44.4%) |  |  |
| Strongly agree | 21 (30.9%) | 44 (59.5%) | 65 (45.8%) |  |  |
| **Patient reported satisfaction** |  |  |  |  |  |
| **I would recommend the approach used during my visit to other patients like me., n (%)** |  |  |  | 0.84<sup>1</sup> | 0.03 |
| Strongly disagree | 2 (3.0%) | 2 (2.7%) | 4 (2.8%) |  |  |
| Disagree | 0 (0.0%) | 1 (1.4%) | 1 (0.7%) |  |  |
| Agree | 7 (10.4%) | 8 (10.8%) | 15 (10.6%) |  |  |
| Strongly agree | 58 (86.6%) | 63 (85.1%) | 121 (85.8%) |  |  |
| Missing | 2 | 0 | 2 |  |  |
| **Did the clinician explain things in a way that was easy to understand?, n (%)** |  |  |  | 0.26<sup>1</sup> | ~ |
| Yes somewhat | 1 (1.5%) | 0 (0.0%) | 1 (0.7%) |  |  |
| Yes definitely | 67 (98.5%) | 74 (100.0%) | 141 (99.3%) |  |  |
| Missing | 1 | 0 | 1 |  |  |
| **Did the clinician listen carefully to you?, n (%)** |  |  |  | 0.58<sup>1</sup> | ~ |
| Yes somewhat | 1 (1.5%) | 2 (2.7%) | 3 (2.1%) |  |  |
| Yes definitely | 67 (98.5%) | 71 (97.3%) | 138 (97.9%) |  |  |
| Missing | 1 | 1 | 2 |  |  |
| **Did the clinician show respect for what you had to say?, n (%)** |  |  |  | 0.95<sup>1</sup> | ~ |
| Yes somewhat | 1 (1.5%) | 1 (1.4%) | 2 (1.4%) |  |  |
| Yes definitely | 67 (98.5%) | 73 (98.6%) | 140 (98.6%) |  |  |
| Missing | 1 | 0 | 1 |  |  |
| <sup>1</sup> Rao-Scott Chi-Square p-value; <sup>2</sup> Missing Value N=1 from Usual Care |  |  |  |  |  |

### Interactions

There were no significant interactions between the intervention and pre-specified patient factors (age, sex, health literacy, and language) for the outcomes of IIRS and A1c.

## Discussion

In this randomized clinical trial conducted at a federally qualified health center, the use of QBSafe cards during routine primary care encounters did not significantly affect measures of fit of care, namely illness intrusiveness, treatment burden, or diabetes distress. Improvements in HbA1c levels were observed in both study arms, with no difference between the trial arms. Nevertheless, the use of QBSafe cards was associated with greater clinician satisfaction with the encounters and improved adherence to diabetes medications among patients, and the QBSafe approach was rated positively by both patients and clinicians.

People living with diabetes face a substantial self-management workload. One study estimated that full adherence to diabetes self-care guidelines would require nearly four hours per day, which is an unrealistic expectation for most, particularly those experiencing adverse social determinants of health.^2^ Our study population, drawn from a federally qualified health center, reflects these challenges: most lived below 150% of the federal poverty level, 37% were insured by Medicaid, and a quarter were uninsured. Participants reported high levels of illness intrusiveness that were comparable to those experienced by patients with cancer,^3^ along with high diabetes distress and treatment burden. Over 40% of patients in the study had a baseline HbA1c level of 10% or above reflecting severe hyperglycemia. These findings underscore the urgent need for interventions that help achieve care that fits and can therefore be feasibly implemented and sustained in the broader context of their life, particularly for patients living with constrained resources.

To date, most interventions aiming to improve fit of care have focused on enhancing patient capacity, often through cognitive behavioral approaches to improve self-efficacy or problem-solving.^31,32^ However, few tools exist to help clinicians reduce the treatment workload they place on patients or to guide conversations on tailoring care to individual life circumstances. Our study suggests that while agenda-setting tools may help initiate such conversations, they may need to be paired with clinician training or system-level support to lead to measurable improvements in care fit and outcomes.

Diabetes care has evolved rapidly over the past decade, with an expanding array of pharmacologic options, new technologies for monitoring and insulin delivery, and increasingly comprehensive and multifaceted clinical guidelines. While these advances offer substantial benefits, they have also made diabetes management more complex, often increasing the cognitive, emotional, and logistical burden on patients. Despite this complexity, relatively little attention has been paid to reducing the overall workload required of patients or supporting care that aligns with their daily lives.^32^ This is particularly important for patients experiencing adverse social determinants of health, for whom treatment burden can directly undermine clinical outcomes. To achieve greater equity in diabetes care, future interventions must prioritize not only enhancing patient capacity, but also simplifying care, supporting patient-clinician collaboration, and designing systems that recognize and respond to the lived realities of patients.

Our findings contribute to a growing evidence base supporting the potential of agenda-setting tools to foster more personalized and collaborative communication in diabetes care. Similar to our study, a Danish trial tested a set of seven conversation cards with patients with type 2 diabetes during routine appointments with diabetes nurses.^33,34^ While the use of cards did not lead to significant differences in the extent to which patient-prioritized topics were discussed, qualitative data indicated that cards added structure and support for raising concerns not traditionally considered diabetes-related. Together, these findings suggest that conversation cards may serve as a low-burden, scalable tool for supporting person-centered conversations, even if their impact on specific outcomes may be modest without further supports or tools for clinicians to help them respond to patient concerns.

Our study has some limitations. This was a single-center study and thus findings may not be necessarily generalizable to other settings. The QBSafe cards were used only once with each patient in the intervention arm; repeated use across encounters may be more impactful. Although training materials were provided to QBSafe clinicians on how to respond to issues brought up by patients, development of a specific toolkit to address illness intrusiveness, treatment burden, and diabetes distress is clearly needed. Finally, our study was limited by a relatively small sample size designed to provide preliminary estimates of efficacy; larger, multi-site studies are needed to test a future iteration of QBSafe intervention on patient and clinical outcomes.

In summary, while the use of QBSafe cards did not improve specific patient-reported or clinical outcomes in this trial, they offer a low-cost, feasible strategy to prompt conversations about treatment burden in primary care. Given the high levels of illness intrusiveness and distress among patients in this underserved setting, efforts to personalize and de-burden diabetes care remain a priority. Future interventions should focus on both enhancing conversations about treatment burden and equipping clinicians to meaningfully respond, particularly in high-need populations where the need for care that fits is greatest.

## Data Availability

The QBSafe cards are freely available for use. Individual participant data that underlie the results reported in this Article will be available from the corresponding author on reasonable request. Participant video recordings cannot be shared due to confidentiality issues.

https://www.patientrevolution.org/qbsafe

## Contributors

All authors had full access to all the data in the study and had final responsibility for the decision to submit for publication.

## Acknowledgement

This study was funded by NIDDK R01DK129616.

## Declaration of generative AI and AI-assisted technologies in the writing process

During the preparation of this work one author (K.J.L.) used ChatGPT-5 in order to improve readability and clarity of the manuscript. After using this tool, the author reviewed and edited the content as needed and takes full responsibility for the content of the publication.

