## Supplemental for "QBSafe: a Randomized Trial of a Novel Intervention to Improve Care for People Living With Type 2 Diabetes"

*Supplement*

**S.Figure 1**: QBSafe conversation cards using in the intervention arm of the trial. QBSafe cards are available online at: https://patientrevolution.org/qbsafe.

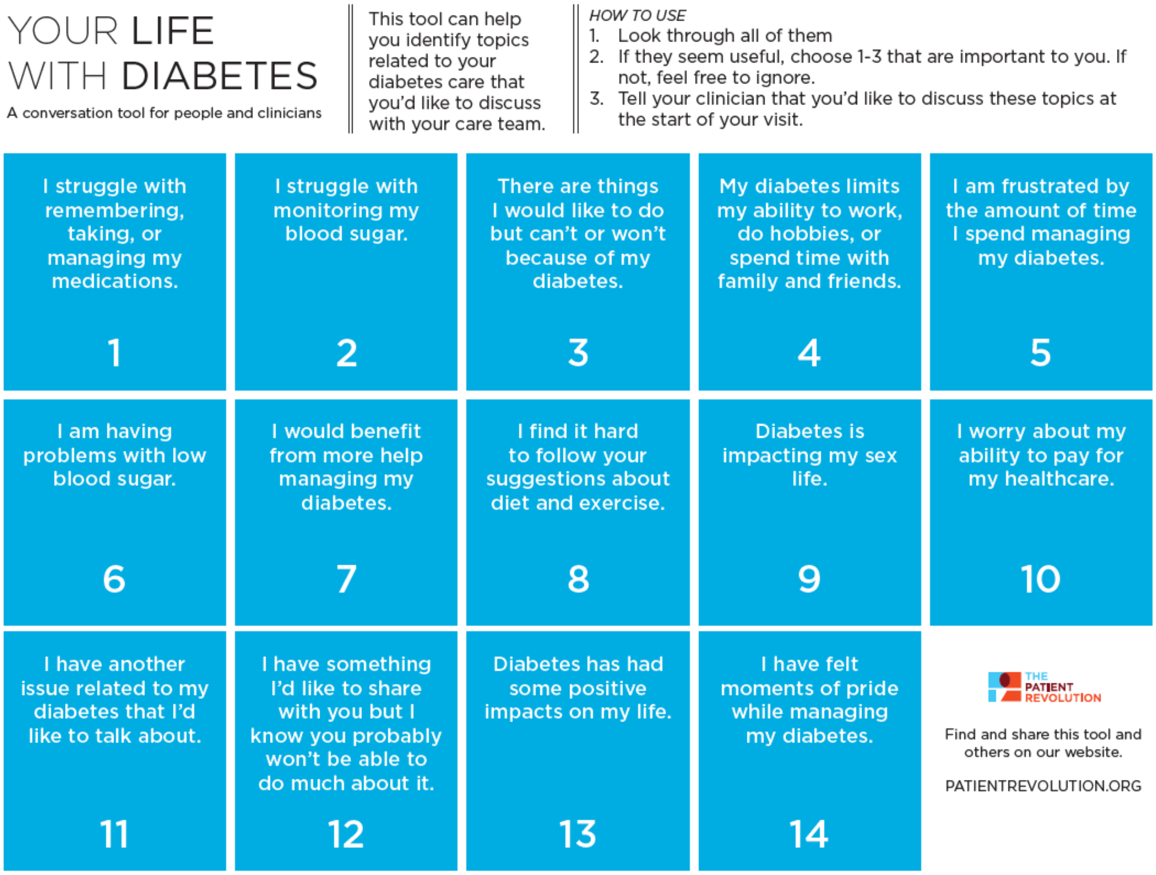

**S.Figure 2A-E**: QBSafe intervention arm materials for clinicians to respond to issues raised by selected QBSafe cards. A. How to use the training materials; B. Responding to treatment burden; C. Addressing sexual health; D. Responding to issues of cost; E. Responding to "pride" cards.

A. How to use the training materials

### QBSafe Clinician Training Materials

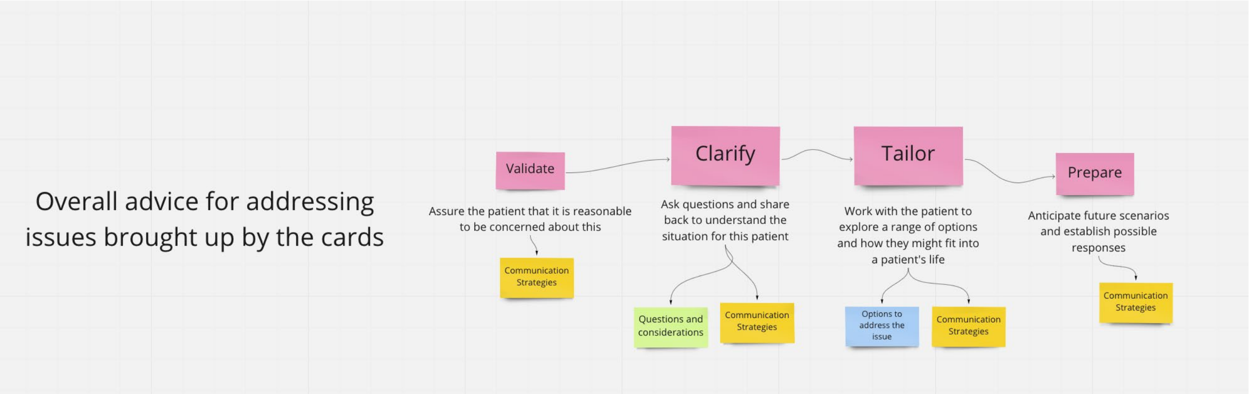


B. Responding to treatment burden

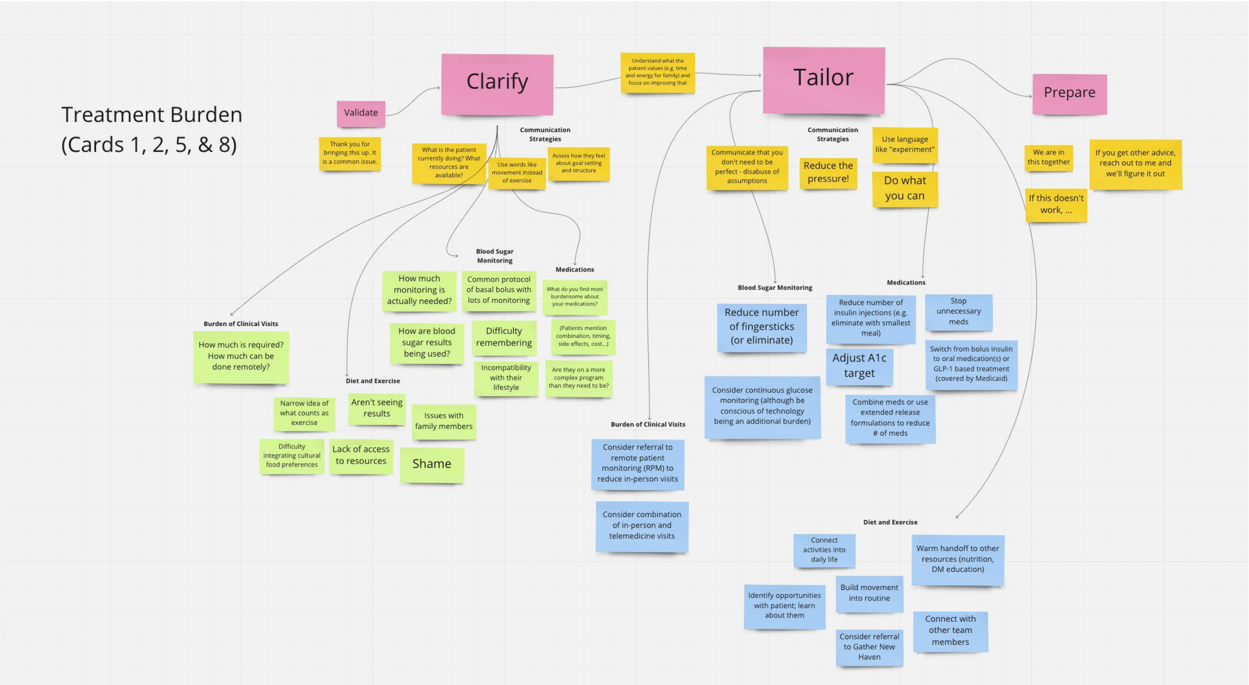

C. Addressing sexual health

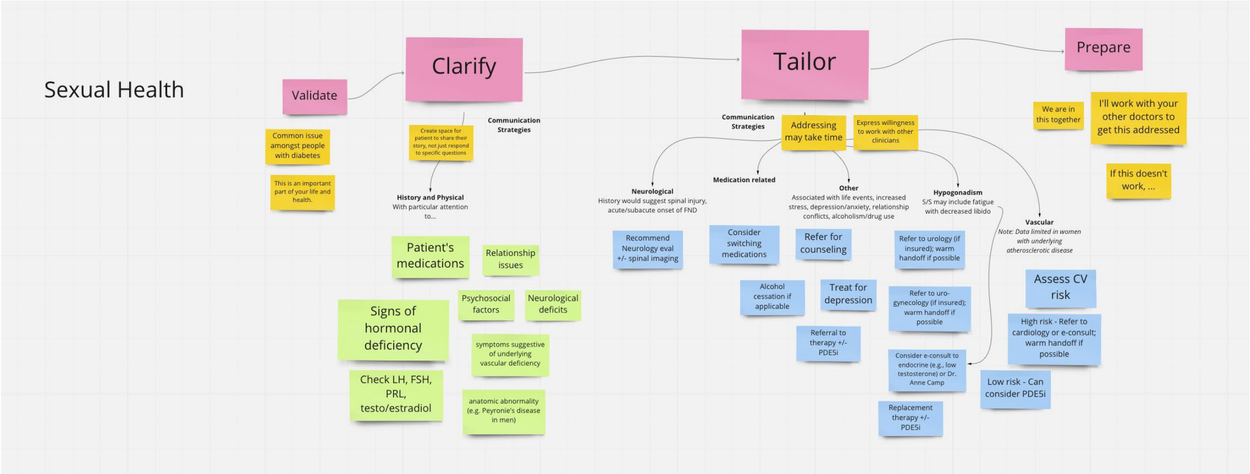


D. Responding to issues of cost

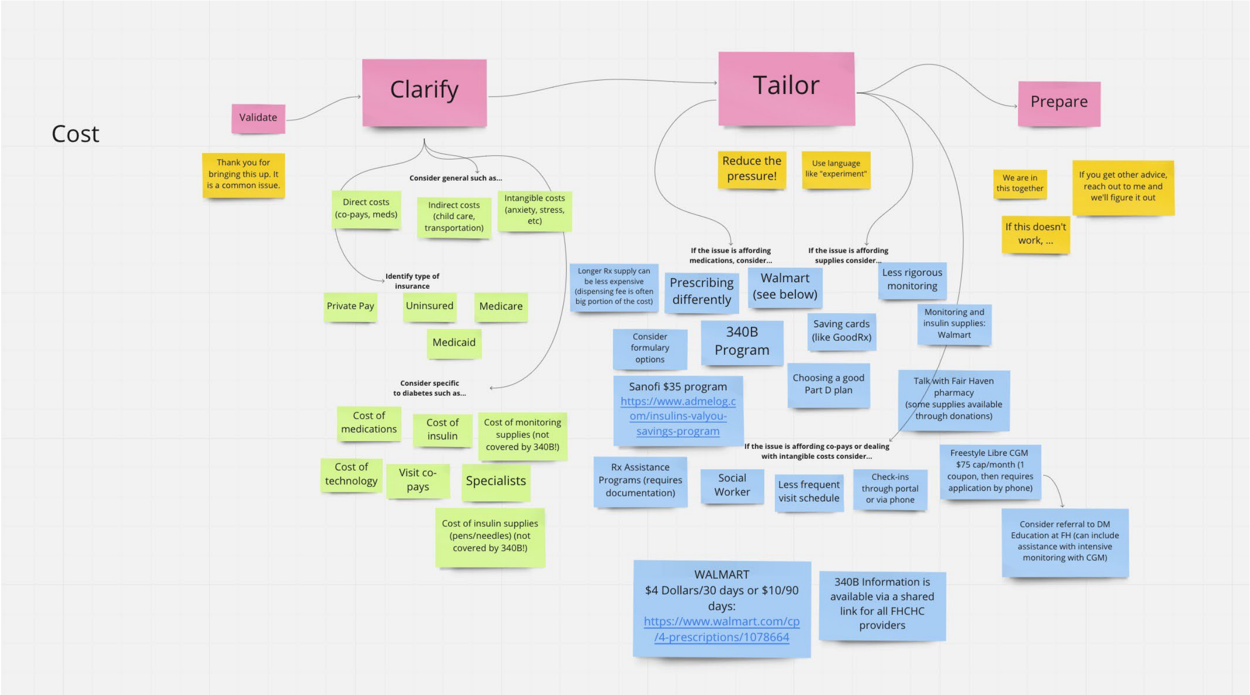

E. Responding to “pride” cards

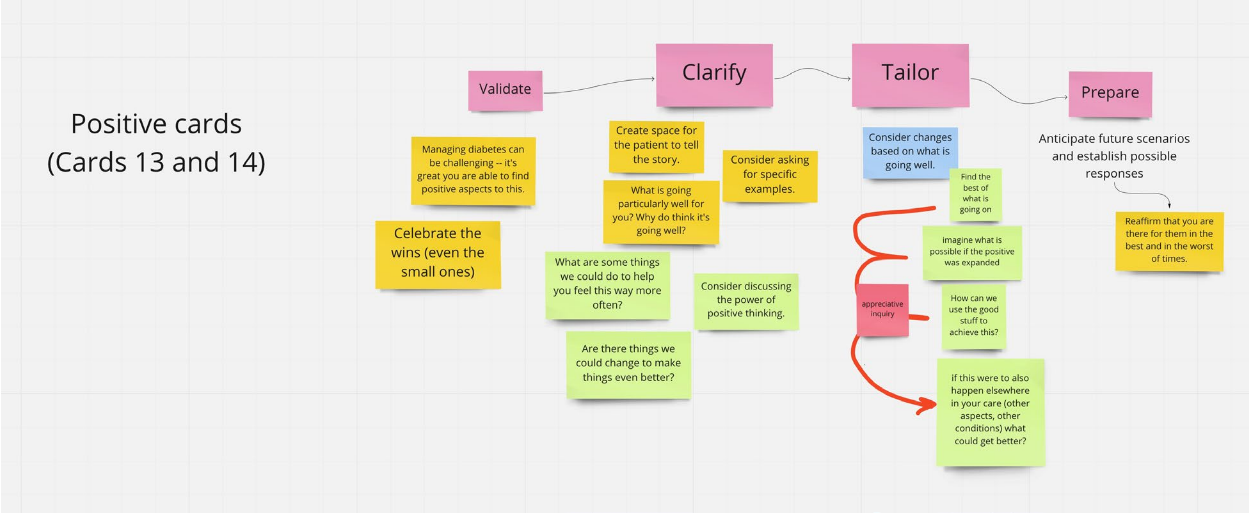

**S.Table 1**: Summary of Diabetes Self-Care Activities (SDSCA) at baseline and 6 months in usual care and QBSafe cards arms.

| | **Arm** | | | **ICC** |
|---|---|---|---|---|
| **SDSCA: Baseline** | Usual Care (N=69) | QBSafe Cards (N=74) | | |
| General Diet | 3.6 (2.57) | 3.8 (2.37) | | 0.01 |
| Specific Diet | 3.8 (1.74) | 3.9 (1.47) | | 0.01 |
| Exercise | 2.8 (2.27) | 2.9 (2.42) | | 0.06 |
| Blood-Glucose Testing | 3.4 (2.80) | 4.1 (2.83) | | 0.002 |
| Foot-Care | 4.1 (2.49) | 3.7 (2.82) | | 0.02 |
| Have smoked a cigarette during the past 7 days | 16 (23.2%) | 18 (24.3%) | | |
| If yes, how many cigarettes did you smoke on an average day? | 8.1 (4.79) | 8.6 (11.51) | | 0.20 |
| **SDSCA: 6-Months[1]** | | | **Mean Difference (95% CI)** | |
| General Diet | 3.8 (2.37) | 3.9 (2.31) | 0.03 (-1.0, 1.0) | 0.05 |
| Specific Diet | 3.8 (1.46) | 4.2 (1.40) | 0.3 (-0.2, 0.9) | 0.01 |
| Exercise | 2.7 (2.07) | 3.1 (2.35) | 0.3 (-0.7, 1.3) | 0.11 |
| Blood-Glucose Testing | 3.6 (2.66) | 4.4 (2.68) | 0.4 (-0.5, 1.3) | 0.004 |
| Foot-Care | 4.4 (2.52) | 4.4 (2.70) | 0.2 (-0.6, 1.0) | 0.03 |
| Have smoked a cigarette during the past 7 days | 15 (26.3%) | 16 (24.2%) | | |
| If yes, how many cigarettes did you smoke on an average day? | 6.6 (5.06) | 6.7 (5.42) | -0.1 (-4.6, 4.4) | 0.37 |

[1]Missing responses for 20 participants (N=12 Usual care, N=8 QBSafe)
[2]Mixed generalized linear model adjusting for a fixed effect of arm and baseline score for the scale, and a random effect of clinician.

**General Diet** = Mean number of days for Q1: Days you have followed a health eating plan; Q2: Have you followed your eating plan.
**Specific Diet** = Mean number of days for Q3: Did you eat 5 or more servings of fruits and vegetables?; Q4: Did you eat high fat foods such as red meat or full-fat dairy products? (Q4 is reversed order)
**Exercise** = Mean number of days for Q5: Number of days your participate in at least 30 minutes of physical activity; Q6: Did you participate in a specific exercise session?
**Blood Sugar Testing** = Mean number of days for Q7: Number of days you tested your blood sugar; Q8: Did you test your blood sugar the number of times recommended by your health care provider?
**Foot care** = Mean number of days for Q9: Did you check your feet?; Q10: Did you inspect the inside of your shoes?
**Smoking** = Average number of cigarettes a day among those that said yes to smoking.
